# Screening for Rheumatic Heart Disease in Asymptomatic Children using Machine Learning from Electrocardiograms

**DOI:** 10.64898/2026.05.03.26352326

**Authors:** Amsalu Tomas Chuma, Chunzhuo Wang, Melkamu Hunegnaw Asmare, Carolina Varon, Jens-Uwe Voigt, Desalew Mekonnen Kassie, Liesl Zühlke, Bart Vanrumste

## Abstract

Early detection of Rheumatic Heart Disease (RHD) is essential in reducing its associated mortality and late complications. In resource-limited settings, automated detection using low-cost electrocardiogram (ECG) sensors can enhance prevention efforts. However, its effectiveness as a potential RHD screening tool in at-risk populations remains unexplored. This study aimed to investigate the utility of machine learning for classifying RHD in a cohort screened for RHD using low-cost ECG devices. The ECGs were collected from 611 at-risk schoolchildren using KardiaMobile, where 47 were confirmed RHD and 564 were healthy. First, the ECG fiducial points were annotated using a publicly available prominence-based delineator. Then, temporal, frequency, wavelet, and visibility graph-based features were extracted from six-leads and fed to the XGBoost classifier. A 10-fold cross-validation was used at different prediction score thresholds to obtain target sensitivity (Se) for screening RHD. Single-lead evaluation on Lead-II showed an F1-score of 60.9%, a Se of 59.6% and a positive-predictive-value (PPV) of 62.2%. However, using multiple leads improved the results, with an F1-score of 62.8%, a Se of 59.6% and a PPV of 66.7%. The best model performance was achieved by adjusting the threshold to 0.6 with Se and PPV of 66% and 51%, respectively. Error analysis revealed that T-wave and STT changes, as well as non-rheumatic mitral valve cases were among the false positive cases. Machine learning can enhance early detection by leveraging relevant ECG features and adjustable target sensitivity based on screening priorities and resource capacity. Measurements can be obtained without chest contact, using only the fingers and knees, thereby enabling use by non-clinical staff. This approach provides a scalable and cost-effective solution for RHD screening in high-prevalence regions.

## I. Background

Rheumatic heart disease (RHD) is a leading cause of cardiovascular morbidity and mortality in children and young adults in low- and middle-income countries (LMICs) [1], [2]. The disease results from recurrent immune cross-reactions to Group A Streptococcus (GAS) during acute rheumatic fever (ARF). These immune responses progressively damage heart valves, leading to the permanent consequence of RHD [1], [3]. Carditis occurs in approximately 50–80% of patients with ARF, but mitral valve prolapse (MVP) has also been reported in association with the inflammatory response before the disease becomes chronic [2]. The cumulative incidence of RHD progression following the first episode of ARF is reported to be 27% at one year, increasing to 44% at five years, and reaching 52% at ten years [2], [3]. This emphasizes the need for early detection of RHD, possibly within ten years after the first ARF incidence.

Echocardiography is the gold standard for RHD diagnosis, as the most important manifestations of RHD are abnormalities of the mitral and aortic valves [4]–[7]. Studies showed hand-held echocardiographic devices (HAND) achieved good sensitivity and specificity, 79% and 87% respectively, in screening for RHD [8], [9]. However, echocardiography-based screening requires equipment and trained personnel, both of which are not readily available in resource-limited settings. Therefore, exploring low-cost and accessible screening mechanisms has the potential to enhance prevention efforts as indicated in [10]. Optional auscultation-based findings in large RHD screening studies reported poor sensitivity and specificity [2], [8]. Compared with an echocardiogram to detect any murmur by a doctor or a nurse had a sensitivity of less than 50%, a specificity of about 75%, and a positive predictive value (PPV) of below 10% [2].

Conversely, machine learning (ML) techniques applied to electrocardiogram (ECG) signals are enabling accurate diagnosis and risk stratification of both structural and functional conditions, including arrhythmias, heart failure, left ventricular systolic dysfunction, hypertrophic cardiomyopathy, and coronary artery disease [11], [12]. Digital biomarkers from ECG intervals, such as QT, QRS, and RR, morphology of QRS and T-wave, and increased QT dispersion support decision-making in clinical practice [12]–[15]. However, some of these markers, such as elevated T-wave or rhythm changes, are considered normal variants [16]. These biomarkers have been suggested as a potential tool to facilitate personalized therapeutic strategies for acquired rheumatic infections [12]– [15], [17]–[19]. Prolonged PR interval, QT prolongation, T wave abnormalities, and elevated TpTe/QT ratio are associated with myocardial and pericardial inflammation. Conduction abnormalities are associated with fibrosis and damage to the sinoatrial node and pathways. Furthermore, atrio-ventricular (AV) blocks and atrial fibrillation have been reported in RHD with a prevalence from 25% to 75% [14].

Extracting the temporal ECG intervals requires defining the onsets, peaks, and offsets of the P, QRS, and T waves (PQRST). These wave fiducial points can be annotated manually by experts or using automated delineators. Manual annotation is a labor-intensive, time-consuming process due to inter- and intraobserver variability, as well as morphological variability across subjects and pathologies [20], [21]. The automation of PQRST annotation has been addressed in a previous communication of our group using various heuristic approaches, including mathematical and wavelet transform (WT) methods, and recently deep learning frameworks [22].

Open-source heuristic-based delineators, such as ECG-kit [20], ECG-deli [23], and neurokit2 [24] have been extensively validated across multiple ECG databases. As reported in [21], [25], [26], heuristic-based methods demonstrate acceptable sample-precision annotation errors comparable with deep learning models, according to the Common Standards for Electrocardiography (CSE) committee’s recommendations [27]. In line with this, in our previous work, we showed the competitiveness of heuristic methods with deep learning approaches on a single-lead ECG dataset recorded from RHD-prevalent asymptomatic schoolchildren [28]. In particular, adopting the method from [25] yielded improved delineation results by incorporating physiology-based interval and segment parameters with visibility graph (VG)-based R-peak detectors to identify an appropriate search window for potential fiducial points. Visibility graphs transform an ECG signal into graph representations to analyze the periodic, random, and fractal properties of the signal [29].

Furthermore, recent studies demonstrate that heuristic-based PQRST delineation is highly reliable in single-lead and wearable ECG recordings. Near-perfect R-wave detection sensitivity of 99.82%, and positive-predictive-value of 99.81% was reported in [30] using MIT-BIH and European ST-T and MIT noise stress test databases. These findings were further confirmed through the systematic validation of heuristic-based delineators on smartwatch ECGs, which exhibited performance comparable to that of deep learning models [26]. The delineation performance decreases in low SNR waves, such as for P and T-waves, with about 89-95% sensitivity [26]. Nevertheless, in ECG interval measurements, a single-lead, low-cost AliveCore KardiaMobile ECG device (AliveCore Inc., Mountain View, California, USA), or KM in short, showed high agreement with clinical 12-lead ECGs [31], [32]. The KM has also been reported for diagnosis, resulting in a sensitivity of 95% for AF, 70% for AV block, and 88% for ST-segment depression [33]. The performance was lower for detection of T-wave inversions (56%), wide-QRS tachycardia (60%), and ST-T abnormalities (33%). In another large case study that consisted of 73,861 participants, the KM achieved a sensitivity of 70% for AF detection by employing a convolutional neural network (CNN) [34]. The study of [32] also reported that prolonged QTc can be detected using an upgraded three-lead AliveCore KardiaMobile 6L (KM6L) with an area under the curve (AUC) of 97%. In advanced RHD, single-lead ECG signal analysis was able to distinguish RHD from the five other most common cardiovascular diseases in low- and middle-income countries, with an accuracy of 51% [35]. However, the use of automated methods to extract and evaluate such features for screening early RHD in at-risk asymptomatic populations has not yet been investigated.

This study investigates the utility of ML for predicting RHD using features extracted from the KM6L, a low-cost ECG device. We used ML to classify participants as either RHD-positive (PwRHD) or healthy (Normal) based on their ECG recordings. We performed the classification task in two steps. First, the ECG waveform is segmented using an open-source, prominence-based PQRST delineator [25]. Second, temporal features and other features that require fiducial points were extracted based on the resultant fiducial points. Finally, the extracted features were evaluated using classical ML algorithms to classify subjects with PwRHD and healthy subjects.

Therefore, the contributions of this study are twofold. First, it explores the potential of low-cost ECG for screening RHD in high-risk populations where there are limited medical resources. To our knowledge, this study is the first automated early RHD screening attempt from ECG signals. Secondly, it proposes relevant rich feature sets from ECG time-frequency and graph representations. By using these features with classical ML classifiers, this study aims to show the significance of low-cost ECG for RHD detection in resource-constrained regions.

## II. Materials and Methods

### A. RHDECG Dataset

The RHDECG dataset contains ECG recordings of 619 schoolchildren who participated in the RHD screening study from March 2023 to September 2023. The study was approved by the ethics committees of the University Hospitals Leuven, Belgium (regnr B3222022001075) and by a collaborating hospital in Ethiopia, Soddo Christian Hospital (regnr SCH1941015). ECG, and echocardiography data were collected from children at four public schools in rural southern Ethiopia. The schoolchildren were aged 10 to 20 years (mean16.1 ± 2.4 years). Further detail about the entire dataset is explained in [36]. Characteristics of the study population are presented in Table I. The diagnosis of RHD was based on echocardiographic findings according to the WHF 2012 guideline [37] (see Appendix A). The guideline suggests findings as normal and abnormal, where the latter further classified into either borderline or definite disease, indicating disease stages. A total of 49 were diagnosed as RHD positives (26 borderline and 23 definite) and 570 had normal findings.

**TABLE I:** Descriptive summary of RHDECG dataset (Mean ± SD)

| Variables | PwRHD(n=49) | Healthy(n=570) |
| --- | --- | --- |
| Age | 16.7 ± 2.4 | 16.1 ± 2.4 |
| Gender |  |  |
| Female | 30 | 329 |
| Male | 19 | 241 |
| BMI | 19.3 ± 2.7 | 19.5 ± 2.7 |
| Heart rate | 81.0 ± 12.1 | 74.7 ± 13.8 |

The ECG data was recorded using KM6L. KM6L is a small pocket device with three electrodes: two on the front side, for the left and right hands, and a reference electrode on the back side, held against the skin on left leg. The three electrodes generate all six ECG channels (I, II, III, aVR. aVL, and aVF) of the Einthoven triangle. The sampling rate is 300Hz. The device is connected via Bluetooth to the “kardia app” from AliveCor Inc., installed on a smartphone (as shown in Fig. 1). The recordings were later transferred to a computer for further analysis. Each recording has a duration of 30-second. The recording physician verified recordings for motion and contact artifacts immediately to allow a new recording if needed.

**Fig. 1:**
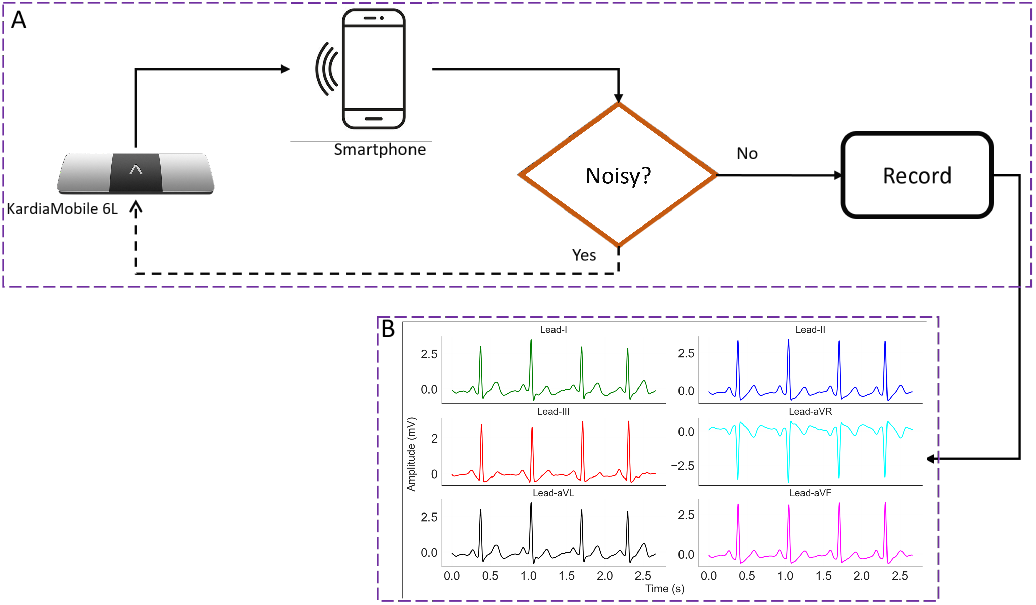
Connectivity block diagram of KM6L and smartphone to record ECG signals. In panel A, first the KM device is connected to a smartphone via Bluetooth that has the Kardia app installed. Then, participants place their fingers on the leads according to KM connectivity manual. The recording physician reviews the real-time ECG signal for artifacts, repeating the recording if necessary for subsequent analysis. Finally, the extracted six-lead ECGs are illustrated in panel B.

### B. Workflow

Fig. 2 illustrates the workflow diagram outlining the overall classification process. The process begins with filtering of the signals, which will be described in section II-C. Next, an open-source heuristic-based delineation algorithm locates PQRST fiducials from the filtered ECG waveforms. Temporal features extracted from the delineated PQRST boundary points, as well as frequency-domain, VG-based, and wavelet features from the input ECG waveforms, will be described in section II-D. Finally, classical ML models were used to evaluate classification performance in 10-fold stratified cross-validation.

**Fig. 2:**
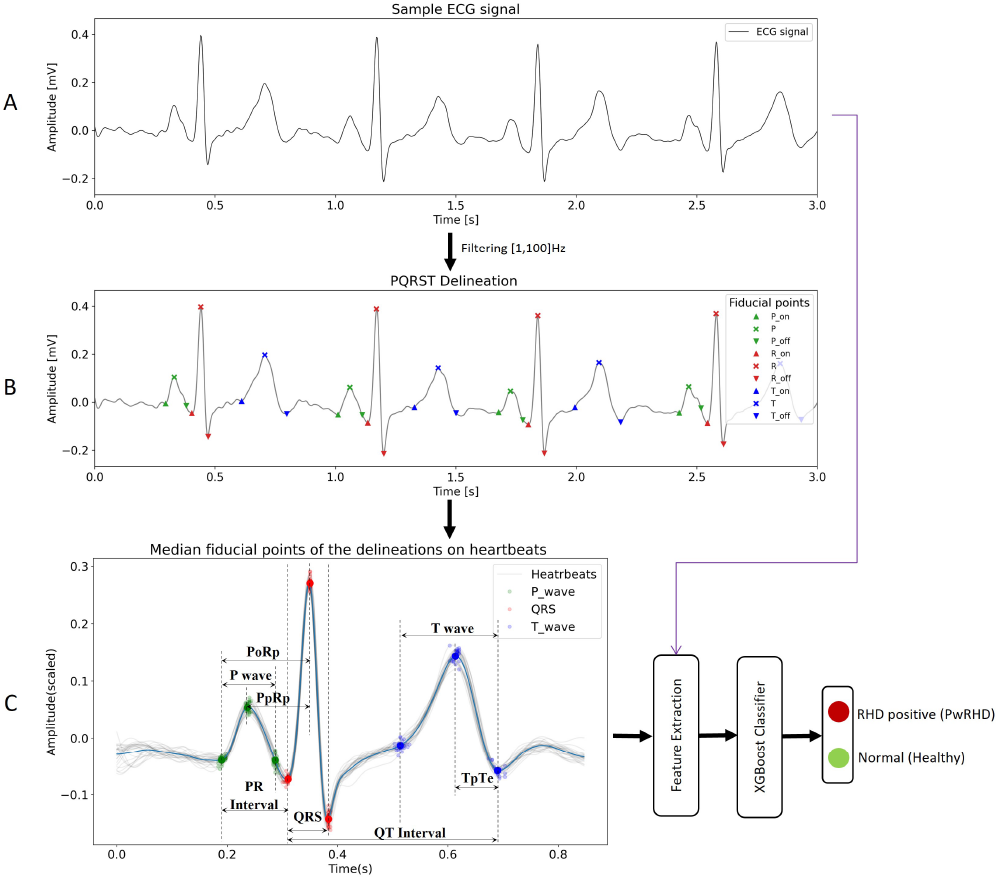
The overall classification workflow diagram. Panel A shows the zoomed-in 3-second single-lead input ECG, and its corresponding delineated PQRST fiducials are shown in panel B Then, panel C depicts overlapped individual heartbeats (light gray lines) centered on detected R-peaks, with the averaged beat ECG shown in a blue line. Key ECG intervals are shown using dashed vertical lines at the median points of each fiducial.

### C. PQRST delineation

#### 1) Preprocessing RHDECG

The RHDECG preprocessing and signal quality assessment was performed following our previous work in [22]. The ECG recordings were first filtered using a zero-phase Butterworth bandpass filter with cutoff frequencies [1-100] Hz to remove baseline wander and high-frequency noise. The signal quality of each ECG using the algorithm proposed by [38]. The algorithm assigns a signal quality index (SQI) value between 0 and 1, where 0 indicates poor quality and 1 indicates best quality of the signal. Then the ECGs with an SQI below 2% percentile in addition to visual evaluation (see Appendix B), which contained two PwRHD subjects and six healthy subjects, were excluded. As a result, these eight recordings were excluded resulting in 611 subjects for evaluation. Finally, following our previous work in [28], we used manual annotations of these ECGs for performance comparison and computed features from the first 15-second window of each ECG for the subsequent classification task based on both automatic and manual annotations.

#### 2) Delineation Method

As we showed in [22], we use the best performing heuristic-based Prominence method by [25] for PQRST delineation in the RHDECG dataset. The 1D U-Net model variants also performed quite well for a similar task [39]. However, they require ground truth manual annotations of all available leads for training. Since heuristic methods do not require manual annotations, they are well suited for automatic delineation of all six leads, as shown in Fig. 3. The delineated PQRST fiducials for these leads were used for analysis. Subsequently, the ECG waveform was split into individual beats, centering on the detected R-peaks. And the median positions of the PQRST fiducial points per each heartbeat were computed for each subject, with all beats aligned to their corresponding R-peaks [40]. Based on these median points, the key temporal parameters, illustrated in Fig. 2 panel (C), were derived.

**Fig. 3:**
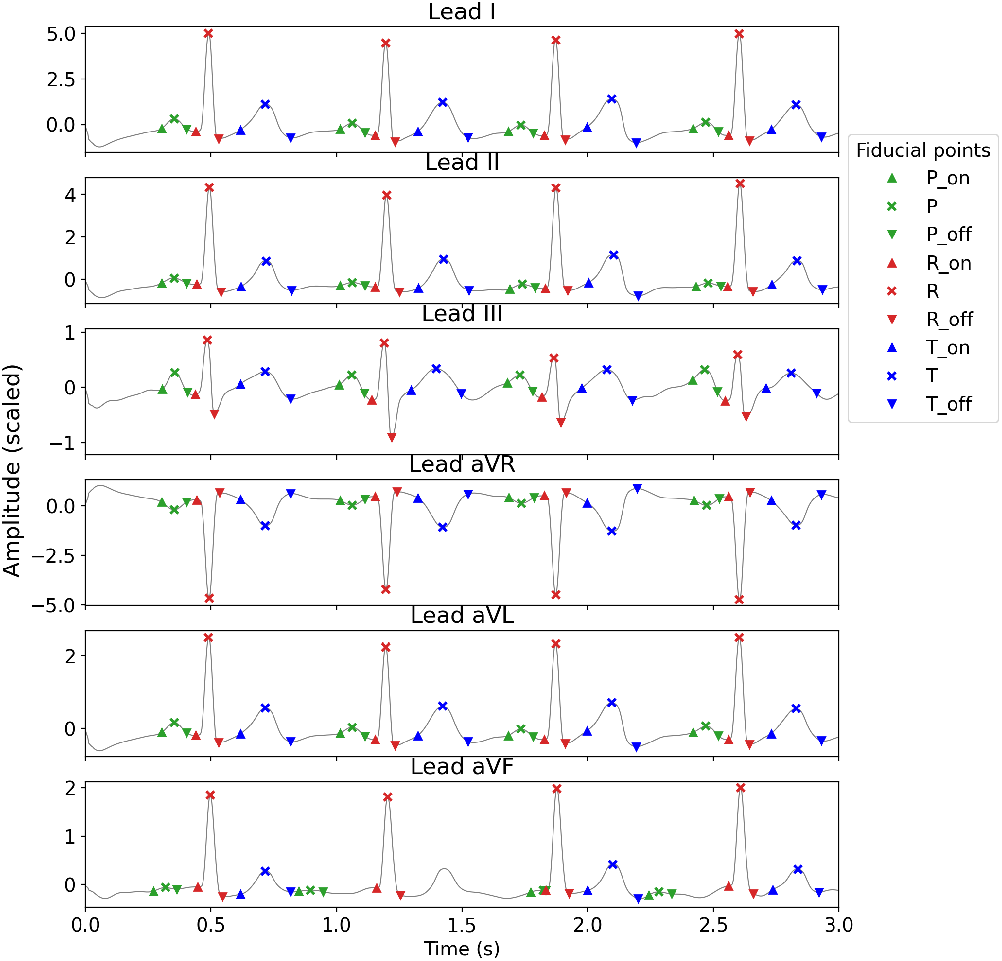
PQRST delineation for an example ECG from RHDECG. PQRST delineation results of an example ECG showing key waves (P, QRS, T) using Prominence method [25]. The markers indicate delineated outputs. Color codes: the P-wave is represented in green colors, QRS in red colors, and T-wave in blue colors.

### D. Classification task

#### 1) Feature extraction

Following the PQRST delineation task, a list of features summarized in Table II was extracted. Features from the temporal-domain, frequency-domain, wavelet-based, and VG-based were extracted from the filtered ECG waveform of each lead. The ECG intervals, area under the PQRST waves, wave durations, and the ratios of these intervals are calculated as follows. For a given beat that contains onset and offset wave boundaries of P-wave (*P*_*on*_, *P*_*off*_), QRS-complex (*R*_*on*_, *R*_*off*_), and T-wave (*T*_*on*_, *T*_*off*_), the intervals were determined as the time between the corresponding onset and offset of the wave. Wave peaks were determined by the sample with the largest amplitude within the wave boundary of the averaged ECG heartbeat signal (see Fig. 3). The areas under the P, QRS, and T -waves were computed as well according to [41].

**TABLE II:** Summary of features extracted from delineated ECG.

| Feature group | List of features |
| --- | --- |
| Intervals | PR, QT, QTc, STT, PT, TpTe, PpRp, PoRp |
| Wave Durations | Pw, Tw, QRSw |
| Ratios | Pw/QRSw, Pw/PR, iCEBc, P-termforce, T-termforce, TpTe/QTc |
| Areas under ECG waves | P-area, T-area, QRS-area |
| Frequency features | pSQI, basSQI, hfSQI, purSQI, rsdSQI, pSig, fSig, zerocrossing |
| Temporal correlations | QRS correlation, Linear Predictive Coding |
| Wavelet features | Wavelet energies per bands |
| Graph features | Density, Degree entropy, Assortativity, Average path |

PpRp: P-wave peak to R-peak, PoRp: $P_{on}$ to R-peak, Pw: P wave, Tw: T wave, QRSw: QRS complex, std: standard deviation

The QTc was calculated according to Bazett’s formula [42]. The PR is the interval between *P*_*on*_ and *QRS*_*off*_, and STT is from *T*_*on*_ to *T*_*off*_. The TpTe interval was measured from the T-wave peak to the T-wave offset. P-termforce and T-termforce are calculated for each lead by multiplying the amplitude at the peak of the wave by the time duration from the peak to the offset of the wave. A peak-to-peak duration of P-wave and QRS (PpRp) was considered from the peak of the P-wave to the R peak. Similarly, the time from the onset of the P-wave to the R peak (PoRp) was also measured. The index of the cardio-electrophysiological balance (iCEB) was calculated as the QT interval divided by the QRS duration 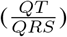. Finally, the wave durations of the P-wave (Pw), the QRS complex (QRSw), the T-wave (Tw), autocorrelation of a beat with the average beat signal, and linear predictive coding (LPC) coefficients [43] were calculated.

In the frequency domain, signal quality index related features were computed, including the relative power in the QRS complex (pSQI), relative power in the baseline (basSQI), relative amplitude of high-frequency noise (hfSQI), relative standard deviation of QRS complex (rsdSQI), signal purity (purSQI), total signal energy (pSig), spectral flatness (fSig), relative P-wave, QRS-wave, T-wave spectra, and zerocrossing rate, as described in [38], [44], [45]. The P, QRS, and T wave bands were [5,30] Hz, [15,40] Hz, and [1,10] Hz as mentioned in [45]. The statistical features, including skewness, kurtosis, linear predictive coding (LPC) coefficients, Singular value decomposition entropy, and the autocorrelation of a beat with the average beat signal were also calculated [45]. Wavelet features were extracted using the discrete wavelet transform with the db4 wavelet at six levels. The estimate of energy at the decomposition frequency bands were calculated [46].

The VG-based features, using both natural and horizontal visibility graphs, were extracted to quantify various properties of the ECG signal’s graph representations [29], [48], [49]. The VG-based features include the density, entropy, assortativity, average path, fractals, and diagonal-based power of scale-freeness (PSVG) of the graph representations of the ECG signal [29], [48], [49]. The VG-based features were computed from the input ECG, a non-overlapping window of 5-seconds, and the downsampled ECG waveforms by factors of 2 and 4. Additionally, metadata of age, gender and BMI were included into feature sets. The participants’ ages ranged from 10 to 20 years, and their BMI ranged from 14.8 to 29.0, thereby both variables were scaled to the [0, 1] range. In total, 373 features were extracted for each input ECG lead.

A supervised binary classification problem was then formulated, where the objective of the classifier *f*_*θ*_(·) is to learn a discriminative mapping from the feature space to the posterior class probability ŷ_*i*_ = *f*_*θ*_(*X*_*i*_). The ground-truth label *y*_*i*_ ∈ {0, 1} denotes the healthy and PwRHD classes, respectively. In case of multilead input, the mean class probability across channels was computed. The eXtreme Gradient Boosting (XGBoost) was used for classification due to its strong performance across a wide range of machine learning tasks [50]. Hyperparameters for the classifier were tuned using Keras GridSearchCV with the search grid values listed in Appendix C. The optimal parameter configurations were subsequently evaluated using 10-fold stratified group cross-validation based on F1-score.

The performance of the classifiers was assessed using subject-stratified 10-fold cross-validation. Evaluation metrics included sensitivity 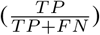, PPV 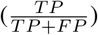, specificity 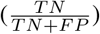, and F1-score 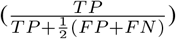. Reported results reflect the model that yields best F1-scores. In addition, to simulate different screening scenarios, the metrics were calculated at a fixed sensitivity thresholds ranging from 0.5 to 0.8 across folds. This allows for adjusting the trade-off between the sensitivity for detecting active cases and the availability of medical resources to perform confirmatory echocardiography. As the RHD diagnostic guideline [5] suggests, such flexibility is useful in disease-endemic areas to focus on the volume of the most likely cases, considering the clinical context and risk factors of the population.

Therefore, three experiments were conducted. The first experiment demonstrates the classification performance obtained using a single lead (Lead-II) based on both manual and automatic PQRST delineations. The second experiment evaluates performance using multiple leads with early and late feature fusion. In early-fusion, features from all leads were combined as input, whereas in late-fusion the class probability outputs from each channel were averaged to obtain the final prediction. The third experiment shows the performance of the best performed model at different sensitivity thresholds.

## III. Results

### A. Experiment on manual vs automatic delineations in single lead ECG

The aim of this experiment is to evaluate comparative performance on the features extracted based on the proposed automated approach and the manual annotations on lead-II. A 10-fold cross-validation classification result for ECGs from PwRHD and healthy subjects using an XGBoost classifier is provided in Table III. The classification result with the manual annotation and automatic annotation showed no significant differences. A slightly better overall detection efficiency was observed using automatic approach, with a higher PPV (61.4% vs 60.9%) and F1-score (60.1% vs 58.2%).

**TABLE III:**
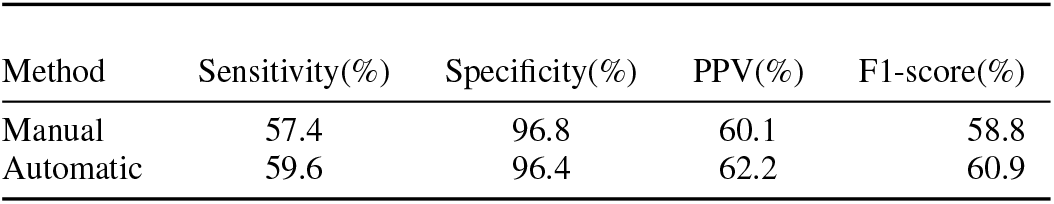
10-fold classification results on Lead-II for manual versus automatic pipelines using XGBoost classifier tuned for best F1-score.

### B. Experiment on automatic delineation using multilead ECG

In this experiment the performance obtained using features extracted from multilead ECGs based on automatic annotations were described. The impact of early and late fusion methods on the classification performance was also assessed. Table IV shows the results obtained with multilead ECGs for both fusion methods. In the early-fusion, the best results yielded by the classifier was an F1-score of 61.1%. The late-fusion approach resulted in a slightly improved F1-score of 62.3%. Particularly the PPV obtained with the late-fusion showed better performance than early-fusion (66.8% vs 62.9%). Which indicates about 33.2% of those flagged as RHD are FP with the late-fusion, while early-fusion resulted in 37.4% of the corresponding FP cases.

**TABLE IV:** Performance comparison of early versus late fusion on multilead ECGs using an XGBoost classifier optimized for best F1-score.

| Method | Sensitivity(%) | Specificity(%) | PPV(%) | F1-score(%) |
| --- | --- | --- | --- | --- |
| Early fusion | 59.6 | 97.3 | 63.6 | 61.5 |
| Late fusion | 59.6 | 97.5 | 66.7 | 62.8 |

### C. Experiment on different target sensitivities using multilead ECG

This experiment aimed at evaluating the best-performing model, described in Section III-B, by adjusting the decision threshold on the class prediction scores to achieve different target sensitivities. Fig. 4 shows the precision-recall (PR) results obtained using the late-fusion approach on multilead ECG ensembles at different sensitivity targets based on class probability score thresholds. Compared with Lead-II alone (see Table III), the multilead ensembles achieved improved performance. At a threshold value of 0.5, the classifier showed an F1-score of 58.1%, a sensitivity of 65.9 and a PPV of 51.7%. A best improvement was observed between 0.5-0.6 as reported in Table IV. At 0.6 threshold, a sensitivity of 51.1%, a PPV of 66.7% and an F1-score of 57.8%. At higher thresholds ranging from 0.7-0.8 the performance drops sharply from a sensitivity of 25.5% at 0.7 threshold to a sensitivity of 14.9% at 0.8 threshold. The corresponding PPV values at thresholds 0.7 and 0.8 were 67.4% and 78.2%, respectively.

**Fig. 4:**
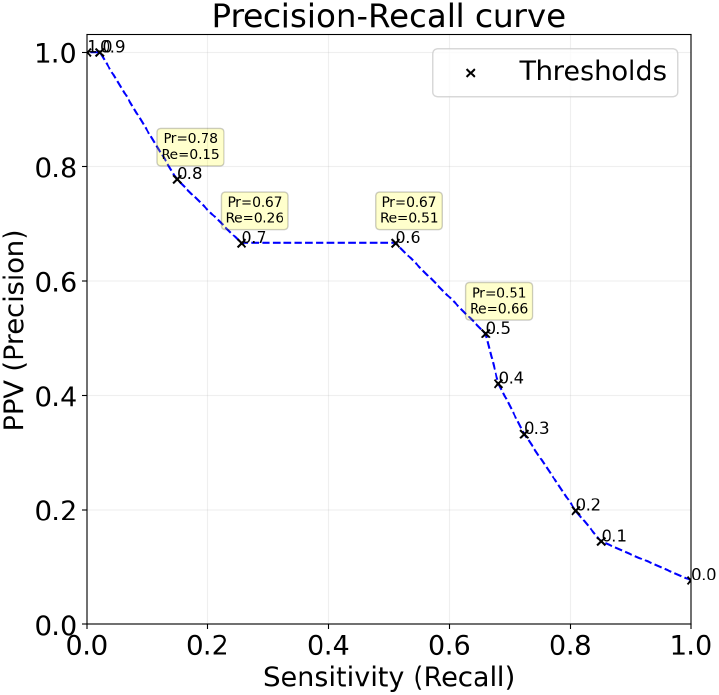
Precision-Recall (PR) curve of the classifier on multilead ECGs at different class probability thresholds. The resulting Se, PPV at each thresholds are annotated by the yellow stickers, where Re: recall and Pr: precision.

Moreover, the contributions of different lead combinations is provided in Appendix D. Although no single lead consistently performed best across all thresholds, leads I, III, and aVL performed relatively better at higher target sensitivities (above 60%). Combining multiple leads in an ensemble further improved performance compared with using any single lead alone.

When analyzing the performance based on disease stage, “Borderline RHD” and “Definite RHD” in test splits is shown at Table V. Overall, better results were observed for Borderline

**TABLE V:** Comparison of classification performance on Borderline RHD and Definite RHD against healthy subjects using multilead ECGs (mean±std)

| Threshold | Borderline RHD |  |  |  | Definite RHD |  |  |  |
| --- | --- | --- | --- | --- | --- | --- | --- | --- |
|  | Sen | Spe | PPV | F1 | Sen | Spe | PPV | F1 |
| 0.5 | 58.7 | 96.4 | 54.5 | 56.9 | 56.4 | 96.8 | 37.9 | 45.5 |
| 0.6 | 53.9 | 97.3 | 62.8 | 57.9 | 58.3 | 92.2 | 31.0 | 46.2 |
| 0.7 | 41.4 | 98.9 | 65.3 | 50.8 | 52.0 | 78.5 | 11.0 | 26.3 |
| 0.8 | 30.2 | 99.3 | 70.1 | 42.6 | 42.6 | 59.6 | 7.3 | 19.8 |

All the metric values are in %.

RHD groups than for Definite RHD groups. For instance, at 0.5 prediction score threshold, the F1-score was 53.3% and the PPV was 56.5% for Borderline, compared to an F1-score of 45.5% and PPV of 37.9% for Definite cases. And similar trends were maintained for the other target sensitivities with a marginal gain of about 1-10% for F1-scores of Borderline cases.

## IV. Discussion

This study demonstrates that low-cost ECG devices with machine learning can effectively detect RHD in high-prevalence asymptomatic populations. A two-stage pipeline combining PQRST delineation followed by extraction of various ECG features, trained with XGBoost classifier, achieved clinically relevant performance. With an adjustable class probability thresholds (0.5-0.8), trade-offs between sensitivity and PPV can be achieved to optimize screening strategies. The proposed ECG-based approach enables shifting the RHD screening task to non-clinical staff or school teachers, as measurements can be obtained without chest contact.

Evidence from prior studies highlights persistent underdiagnosis in resource-limited regions due to limited access to echocardiography [8]–[10]. Auscultation-based screening, although more accessible, has limited diagnostic accuracy and inconsistent clinical interpretation [8], [9]. In contrast, the proposed ECG-based automated approaches offer an alternative method that can be implemented at scale with minimal training. This enables screening tasks to be shifted from highly specialized clinicians to community health workers or school personnel, improving access in underserved settings. Such an approach also supports scalable deployment in routine community health programs without requiring advanced clinical expertise or specialized infrastructure.

### A. Screening potential of an ECG in RHD-endemic regions

The findings of this study show minimal difference in performance between the features extracted with an end-to-end automatic approach and manual annotation methods in single-lead ECG. Table III shows that automatic annotations were as good as, or sometimes better than, manual annotations on Lead-II. In multilead analysis, shown in Table IV, the late-fusion approach performed slightly better than the early-fusion approach. This suggests that modeling each ECG lead independently helps the classifier capture lead-specific patterns and reduces the impact of signal noise on prediction. The reason is that different leads can provide complementary information, where cleaner signals in one lead can compensate for noise in another, particularly in ambulatory settings.

ECG-based detection of RHD showed promise as an automated screening tool in resource-limited settings. While detecting early cases is important with higher sensitivity targets, it is necessary to balance the burden at nearby hospitals during confirmatory checkups after the screening. As shown in Fig. 4, at 0.6 prediction score threshold, the model achieved a PPV of 66.7% suggesting that about a one-third (33.3%) of positive predictions does not represent the true RHD cases. Increasing the threshold to 0.8 decreased the sensitivity to 14.9% while elevating the PPV to 78.2%. This may lead to unnecessary clinical referrals, thereby increasing the burden on the healthcare system. In addition, repeated false alarms may cause psychological stress in participants, reduce trust in screening programs, and discourage future medical follow-up. Nevertheless, setting the threshold between 0.5-0.6 showed an optimal results, yielding a sensitivity and PPV of 59.6% and 66.7%, respectively (see Table IV).

Compared with alternative screening modalities, these results are comparable to those of HAND and superior to auscultation-based screening. For example, auscultation-based RHD screening studies [2] reported 38.2% sensitivity, 75.1% specificity, and 5.1% PPV, missing the majority of participants with early RHD [2], [9]. Whereas the ECG-based model, within similar sensitivity target, achieved an improved both specificity and PPV by significant margin. Similar to the auscultation-based outcomes, cases with non-rheumatic abnormalities were also classified as positive. Overall, the ECG-based classifier performed better than auscultation at the evaluated sensitivity targets from 26-67% (see Fig. 4). Therefore, in the context of RHD screening in resource-limited regions, different working points could be required because disease prevalence and available resources vary across risk areas. The PR-curve in Fig. 4 suggests the model performance at a sensitivity of 50–60% for low-to moderate-risk areas to maintain PPV, and *>*60% for high-risk areas to reduce missed cases. This ensures a balance between detection performance, resource constraints, based on the local epidemiological conditions.

Although handheld echocardiography based screening has reported high concordance with portable echocardiography in definite RHD [8], the accuracy for detecting borderline RHD was poor with sensitivity and specificity of 62% and 82%, respectively [7]. Our classifier has also achieved comparable performance with HAND, as shown in Fig. 4. For instance, at 50-60% target sensitivity range, the ECG-based model achieved an average specificity of about 97%, and a PPV of 51-67% (see Fig. 4). Moreover, stratified evaluation by disease severity showed that borderline RHD achieved higher detection performance than definite RHD (see Table V). This observation may be partly explained by more active pathological processes during the early stages of the disease in the study cohort, although it may also be influenced by the limited number of definite RHD cases available for training. Overall, these results indicate that the ECG-based model performs comparably to the previously reported HAND-based approach in [7].

### B. Error Analysis

Further analysis of the confusion matrix extracted from the Fig. 4 at a prediction score threshold of ≥ 0.6 revealed that tall T-waves or ST changes in 11 (F=8, M=3), and non-rheumatic mitral valve abnormalities in 4 (F=3, M=1) cases within FP groups (n=13). Elevated T-waves associated with myocardial electrolyte abnormalities and can be a normal variant in young patients and athletes [16]. ST changes are also due to electrolyte imbalances, although there is no direct evidence associating them with sex or ARF. These observed mitral valve abnormalities were associated with MVP without rheumatic etiology. The association of MVP cases is debated, as it may be a consequence of an inflammatory response in patients with acute rheumatic carditis [53], while some studies report no association between MVP and ARF [51].

The model misclassified a greater proportion of definite RHD than borderline RHD. For instance, a 45% of the definite and 36% of the borderline cases were misclassified at an average sensitivity of 59.6%. In our previous work on symptomatic cases [35], definite structural manifestations were always reflected as detectable electrical signatures with a detection accuracy of 94%. However, our result suggests such structural severity associated with RHD is not always reflected in the ECG features for asymptomatic cases. This reaffirms the non-specificity of ECG changes in subclinical RHD [52]. Nevertheless, at PPV between 51.7 and 66.7%, approximately 51.1-65.7% of true RHD cases were correctly detected (Fig. 4). This is essential for enabling timely intervention by identifying individuals with suspected RHD, particularly given its often silent clinical progression.

### C. Limitations

The main limitation of this study is the small dataset size which perhaps hindered the model’s ability to generalize better. It is worth noting that increasing sensitivity introduces relatively a lower PPV at greater target sensitivities ≥ 75% which raises concerns regarding potential overdiagnosis. This could overwhelm local healthcare centers and may negatively impact individuals who are misdiagnosed to unnecessary psychological stress.

Therefore, further validation on a larger dataset with more RHD cases, and data fusion techniques combining the ECG features with other different physiologic and clinical history data to address the multi-factorial nature of disease is beneficial. This may encompass the use of synchronized ECG and auscultation data, or independent data modalities including echocardiography data. Fine-tuning pretrained foundation models, as described in [54], on multimodal ECG, heart sounds, and electrocardiograph images will be explored in the future study.

## V. Conclusion

ECG can play a vital role in screening asymptomatic children for RHD in highly prevalent regions. At target sensitivity thresholds of 50-80%, an ECG-based model can discriminate children with RHD from their healthy counterparts with an FPR ranging from 51-78%. For an optimal setting that balances sensitivity and PPV trade-off, a moderate prediction score threshold between 0.5-0.6 can be used. In RHD endemic high prevalent regions, prioritizing sensitivity is critical for minimizing missed RHD cases and mitigating long-term risk. However, in resource-limited environments, this trade-off must be balanced to prevent overburdening local healthcare systems.

The proposed approach offers significant value as a low-cost, scalable screening tool for population-based active case surveillance in endemic regions. Most importantly, it allows task-shifting as measurements can be obtained without contact with the chest but only using the fingers and knee. This approach therefore provides an alternative solution for RHD screening without requiring skilled clinical personnel. While echocardiography remains the most sensitive screening method, leveraging low-cost wearable sensors such as ECG can enhance automated RHD detection. This study demonstrates the value of ECG in early RHD identification and underscores the need for further research on a multi-modal approach to refine early detection and management strategies.

## Data Availability

All data produced in the present study are available upon reasonable request to the authors

## VI. Abbreviations

RHD: Rheumatic heart disease
ARF: Acute Rheumatic Fever
ECG: Electrocardiogram
PQRST: P, QRS and T waves
KM6L: KardiaMobile 6L
HAND: Hand-held Echocardiography devices
MVP: Mitral valve prolapse
VG: Visibility graphs
CNN: Convolutional Neural Networks
DL: Deep Learning
ML: Machine Learning
KM6L: Kardia Mobile 6L ECG sensor
DNN: Deep Neural Network

## Acknowledgment

The authors are grateful to the RHDECG dataset collaborators, the study participants, the schools and hospitals involved, and the examining physicians during the study campaign. We also extend our gratitude to the Flanders AI program for supporting this study.

## Appendix

### A. The WHF echocardiographic criteria for RHD diagnosis

**Fig. 5:**
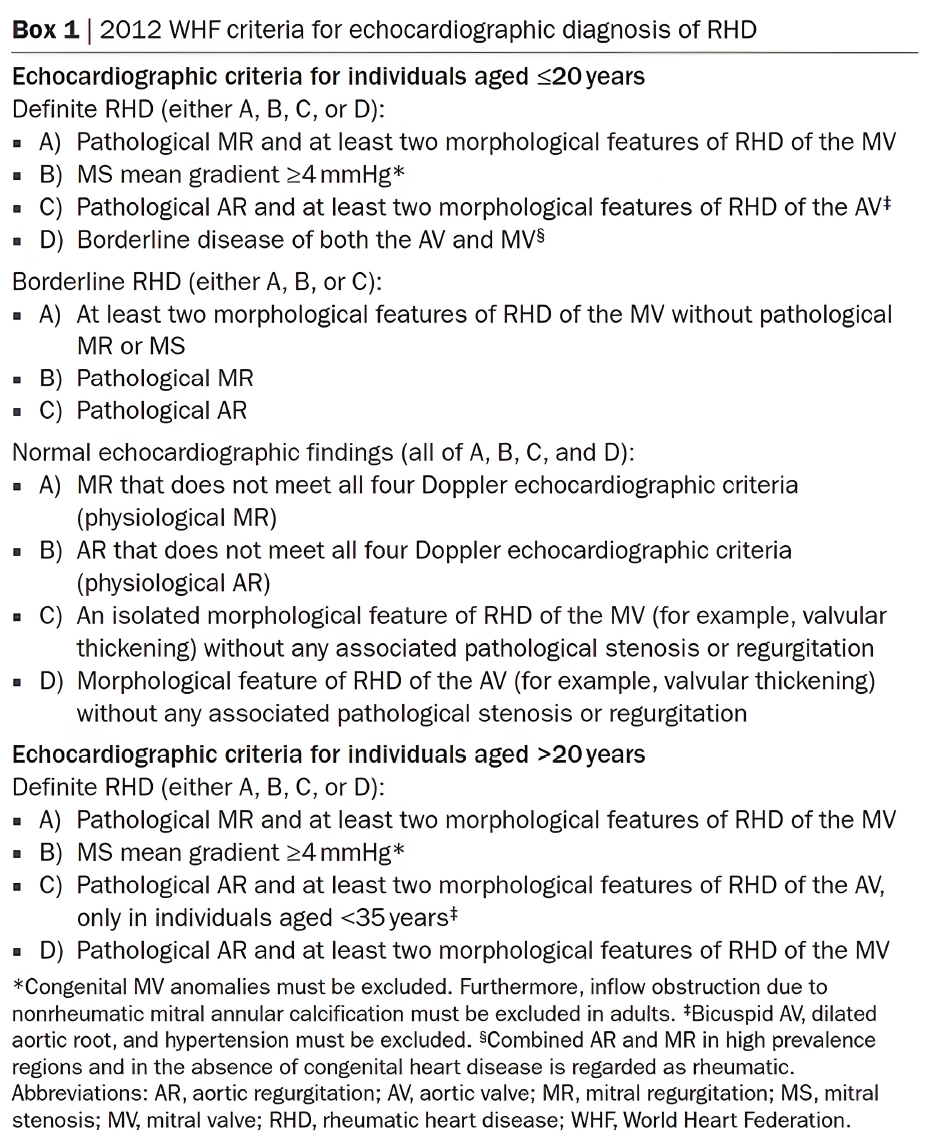
The 2012 WHF echocardiographic criteria for RHD diagnosis from [37].

### B. Signal Quality Assessment

The quality of each ECG signal in the RHDECG dataset was assessed using the algorithm [24]. The values range from [0,1] where 1 indicates a best quality ECG, and 0 indicates the worst quality. In the Fig. 6 shown below the least 2% ECGs were also assessed visually and were excluded from the further analysis in the study.

**Fig. 6:**
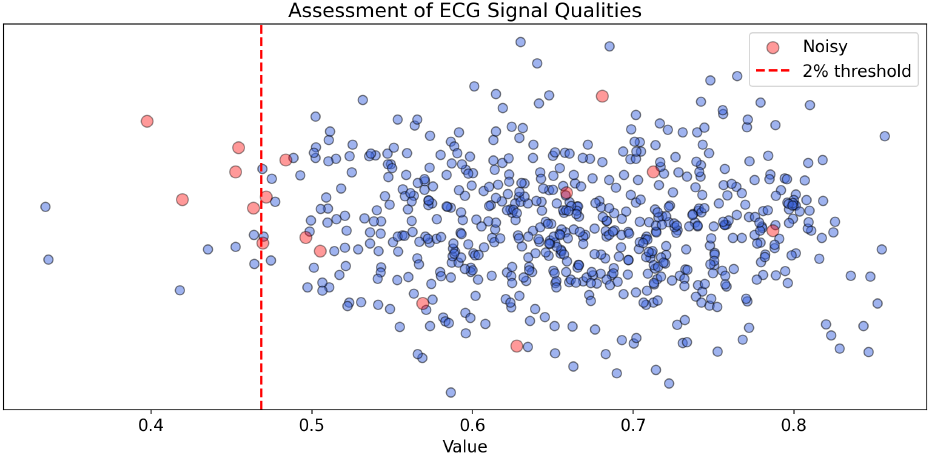
The ECG signal quality assessment based on the results from [24]. The x-axis shows ECG signal quality index values for each input ECG. The colored (orange) data points are those labeled as noisy based on visual evaluation.

### C. Hyperparameters grid table for different ML classifiers

The following table shows the list of grid values used for hyperparameter search for the XGBoost model.

**TABLE VI:** Hyperparameters grid table for different ML classifiers.

| Classifier Model | Hyperparameter | Values grid |
| --- | --- | --- |
| XGBoost | n_estimators | [51,151,251,301] |
|  | learning_rate | [0.001,0.01,0.1] |
|  | max_depth | [3,5,7,10] |
|  | min_child_weight | [1,3,5,7] |
|  | subsample | [0.5,0.7,1.0] |

### D. Evaluation of combinations of the ECG leads for classification of RHD using XGBoost

The leads were combined for all possible combinations of the six-leads (I, II, III, aVR, aVL, aVF). To show the top best contributing combination, we show only subset of the top 5 combinations at each prediction score threshold value. The analysis considers all possible combinations of the six standard ECG leads (I, II, III, aVR, aVL, aVF). To identify the most performed lead configurations, performance is evaluated across all combinations at each decision threshold. For each threshold level, only the top five lead combinations with the highest F1-scores are shown, across the prediction score threshold points.

**Fig. 7:**
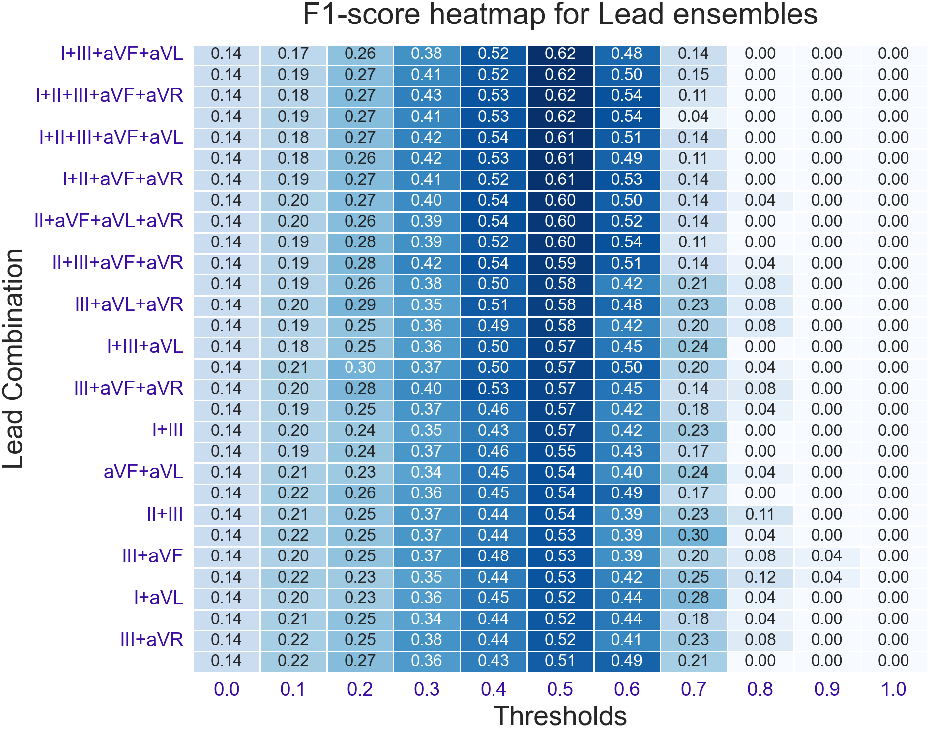
The F1-scores for ensembles of the ECG leads at different sensitivity thresholds. The x-axis indicates sensitivity threshold, and y-axis shows specific ensemble of leads contributions at a corresponding thresholds. The color intensity aligned with the score values.

### E. Features Importance

The feature importance summary plot in Fig. 8 illustrates the top 30 features. The features useful for classification across the entire evaluation set are aggregated and averaged across folds. While the precise contribution of individual features varied across cross-validation folds, features related to T-wave, QRS, P-wave, time-frequency energy at bands [1-5] Hz and [30-45]Hz, relative high-frequency, entropy, PSVG, and fractals consistently ranked among the most influential for prediction across folds. There was also a high contribution of sex, age and BMI features.

**Fig. 8:**
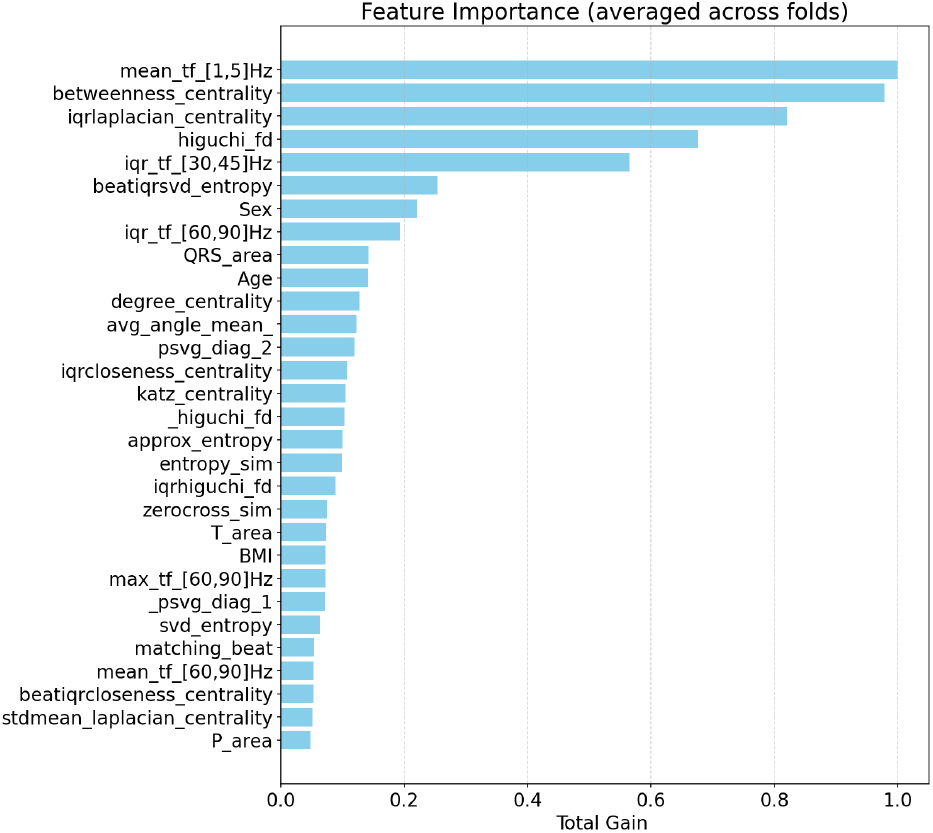
Aggregated feature importance summary plot of top 30 features across the 10-folds. The x-axis shows scaled feature importance values. The y-axis shows list of top-30 important features.

**Fig. 9:**
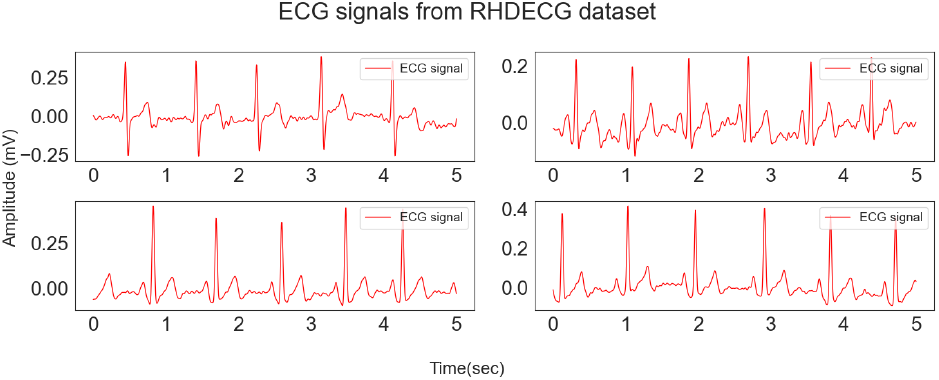
Examples of misclassified PwRHD ECG

The ECG manifestations related to RHD are non-specific [18]. Symptomatic RHD may cause various arrhythmias, such as AF, PVC, and bradycardia, and persistent morphological changes, such as P-mitrale, notched and wide QRS, prolonged PR, and prolonged QTc [18], [52]. Such morphological and rhythmic alterations were not commonly observed in our study cohort (see Fig. 9). In the top-left ECG, the P-wave was not prominently discernible, whereas the top-right tracing displayed tall P-waves. Aside from these features, the remaining tracings predominantly exhibited temporal variations rather than morphological changes of the waves. However, the PR interval, Pw/PR ratio, and P-wave duration variability were reported as strong predictors [28] in early RHD. The feature importance graph shown in Fig. 8 partially reflects that wave related parameters, including the areas under P, T and QRS waves are still important across various cross-validation sets.

Moreover, the energies in the frequency bands related to T-waves, P-wave, and the QRS wave at bands [1-5] Hz, [30,45] Hz, and a high frequency band related to abnormal QRS at [60,90] Hz were also important for the classification.

In PwRHD ECGs, the relative QRS power with respect to the total power is higher in comparison to ECGs from healthy groups [35]. The authors in [38] indicated that the energy of the QRS wave is concentrated in a frequency band centered at 10Hz and is 10Hz in width. However, this frequency bandwidth ranges from 8Hz to 50Hz in a normal ECG, with the upper frequency limit exceeding 70Hz in abnormal sinus rhythms [55]. The VG-based features are also important, particularly adjacency matrix diagonals, its entropy, and centrality. These features were also showed improved results on classification of different cardiac disorders [29]. Similarly, PSVG values for patients with congestive heart failure significantly differed from those that have normal sinus rhythm [48]. Although the PSVG features were not statistically significant in our cohort, their importance was observed for PwRHD classification as well.

